# Patient-centered Evaluation of AI Answers to Genetic Counseling Questions

**DOI:** 10.1101/2025.03.27.25323779

**Authors:** Yidi Huang, Hita Kambhamettu, Elisabeth Wood, Cara Cacioppo, Demetrios Ofidis, Rajia Mim, Angela Bradbury, Kevin Johnson

## Abstract

The growing use of large language models for health communication raises important questions about patient preferences, trust, and satisfaction with AI-generated content. We conducted a qualitative survey comparing patient perceptions of clinician versus AI answers to questions about Alzheimer’s disease and genetics. Twenty-six participants scored responses on relevance, trustworthiness, and coherence, and additionally selected a preferred response and provided free-text comments. We found that participants generally preferred AI responses over human-written ones and rated them higher on all evaluation axes. Qualitative analysis of free-text comments identified key factors influencing patient preferences, including clarity, verbosity, certainty, empathy, and numeracy, with patients showing diverse and sometimes contradictory preferences along these dimensions. These findings suggest the AI-generated responses were well-received by patients and may have a role in triaging patients with information-seeking queries. Future work should focus on dynamically identifying patient communication preferences and tailoring communication styles accordingly.

## Introduction

Patient-centered care has been at the forefront of healthcare for over a decade, with previous work identifying how this critical issue has been a concern for both healthcare organizations and patients^1^. With the rapid advancement of artificial intelligence in healthcare, specifically generative AI^2^, ensuring that AI-assisted systems integrated into health infrastructures do not compromise patient-centered care has become imperative. Incorporating AI within the context of patient-centered care needs to be carefully studied to create patient buy-in and ensure the effective adoption of these technologies. Moreover, choices that promote decision self-efficacy and patient perspectives on this emerging technology should be incorporated so that the use of AI in healthcare is beneficial and equitable for all patients.

AI-enabled healthcare may help to increase equity in health outcomes, reduce diagnostic errors, improve treatment protocols, and offset increasing labor demands and staffing shortages among health practitioners. Recent studies have demonstrated that large language models (LLMs) exhibit promise in various healthcare-related applications, including in interpreting lab test results^3^, genetic test counseling^4,5^, and answering medical questions^6–8^.

Further exploration has focused on refining LLMs’ therapeutic behavior using reinforcement learning from human feedback (RLHF)^9^. A comparative assessment using de-identified patient-physician messages demonstrated that ChatGPT’s responses scored higher on empathy metrics, as evaluated through both algorithmic and human methods^10^. LLMs have also been utilized to improve patient education materials^11,12^, achieving higher readability when evaluated by medical specialists.

Patient perspectives on LLM-generated responses reveal higher satisfaction levels compared to clinician responses in certain contexts, particularly when addressing patient medical advice requests^13^.

We focus on the application of LLMs in a domain where conveying complex health information is critical for shared decision-making between patient and clinician: genetic testing, specifically in testing for risk-modifying variants that reveal an individual’s risk for developing Alzheimer’s Disease (AD). With a demand for genetic testing outpacing the increase in genetic counseling positions, LLMs offer a potential solution to triage patients and support the workplace demands on human counselors.

In this paper, we seek to build upon this suite of prior work and others in order to better understand patient’s attitudes towards AI-generated responses about answers to frequently asked questions about AD and genetics. We report results from a qualitative survey study of 26 US-based adults where participants compared AI-generated and human written responses to questions relevant to AD and genetics. Overall, we find that participants prefer AI-generated answers over human-written answers to questions about AD and genetics. The results are discussed below in the context of patient-centered care in the hopes that more patient and public concerns regarding this technology are incorporated into medical standards.

## Methods

To better understand patient attitudes towards AI-generated content in genetic counseling, we conducted a survey in which respondents were asked to compare human- and AI-generated answers to the same question. Participants were recruited through the Penn Telegenetics program from an existing study cohort who participated in user testing for digital tools for ApoE testing. Participants were offered a $25 gift card as a reward for participation. These activities were approved by the University of Pennsylvania Institutional Review Board.

Each participant was administered a survey consisting of three responses to each of ten frequently-asked questions about AD and ApoE results. Each question was paired with three responses: 1) *Human* - a response written by a genetic counselor; 2) *GPT* - a response generated by simply prompting GPT-4; and 3) *RAG* - a response generated by our previously described retrieval-augmented pipeline^14^, which retrieves relevant context from a bank of recorded counseling sessions. The full set of questions and responses can be found in Supplement S1. Participants were shown each question and all three responses in blind randomized order. For each response, participants were asked to rate their agreement with the following questions on a 1-5 Likert scale: (1) This response is relevant to the question, (2) I trust this response, (3)This response makes sense. We selected these three axes of comparison based on the Patient Education Materials Assessment Tool (PEMAT)^15^.

Finally, participants were asked to provide optional free-text comments on each response and to choose their preferred response.

We performed statistical analysis and plot visualizations of the numerical responses using Python and the Seaborn visualization library^16^. To analyze the free-text comments, we performed a qualitative thematic analysis^17^ of expressed preferences. We conducted an initial code-development phase by inspecting the data both manually as well as using GPT-4-generated summaries. Code definitions were finalized across multiple discussions among the authors. In addition to preference themes, we also annotated mentions of clarity using the codes *clear* (indicating clear responses or other praise for explanation quality) and *vague* (indicating vague or confusing responses or other criticisms of explanation quality). Finally, we manually annotated the comment corpus with our codebook using the Prodigy annotation tool^18^.

## Results

We received 30 responses to the survey, with 26 individuals completing the full survey. Due to a technical error, one question-response set was corrupted and excluded from analysis. The final analysis set consisted of 26 patient responses to nine question-answer sets.

Patients expressed a preference for AI-generated responses over human responses overall. Both GPT and RAG responses received higher average ratings on trustworthiness, relevance, and coherence than human responses (Table 1, Figure 1). Additionally, AI responses were more often selected as preferred than human responses (n=234; GPT=94/0.40; RAG=58/0.25; Human=46/0.20; No preference=36/0.15) (Figure 2). Human responses were also more likely to be described as vague in free-text comments than were RAG and GPT responses (Figure 3).

**Table 1.** Average Likert scores by response source. * indicates significance in T-test comparison with Human at α=0.05

|  | Human | RAG | GPT | AI (RAG \|\| GPT) |
| --- | --- | --- | --- | --- |
| Coherent | 3.88 (1.09) | 3.99 (1.00) | 4.25 (0.92) * | 4.12 (0.97) * |
| Trustworthy | 3.83 (1.06) | 3.97 (1.03) | 4.19 (0.93) * | 4.08 (0.98) * |
| Relevant | 4.04 (0.97) | 4.06 (0.94) | 4.31 (0.86) * | 4.18 (0.91) |

**Figure 1.**
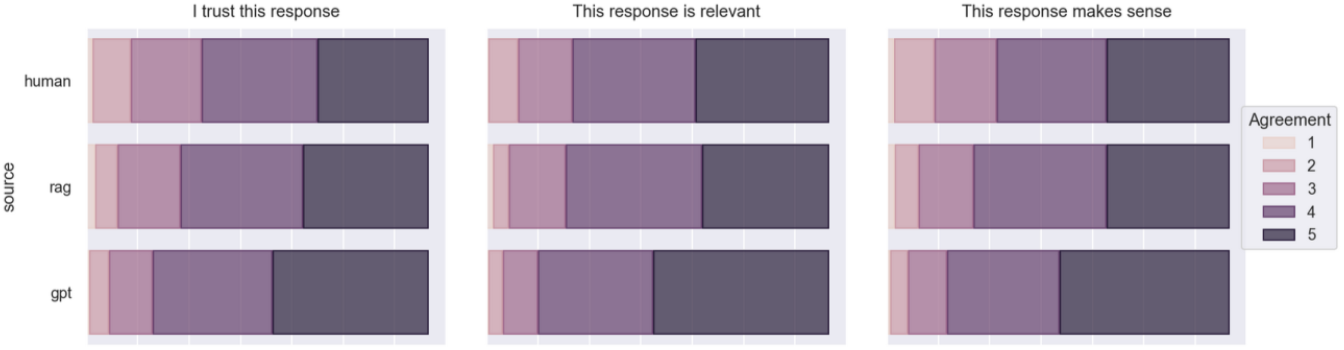
Average numerical ratings among different response sources.

**Figure 2.**
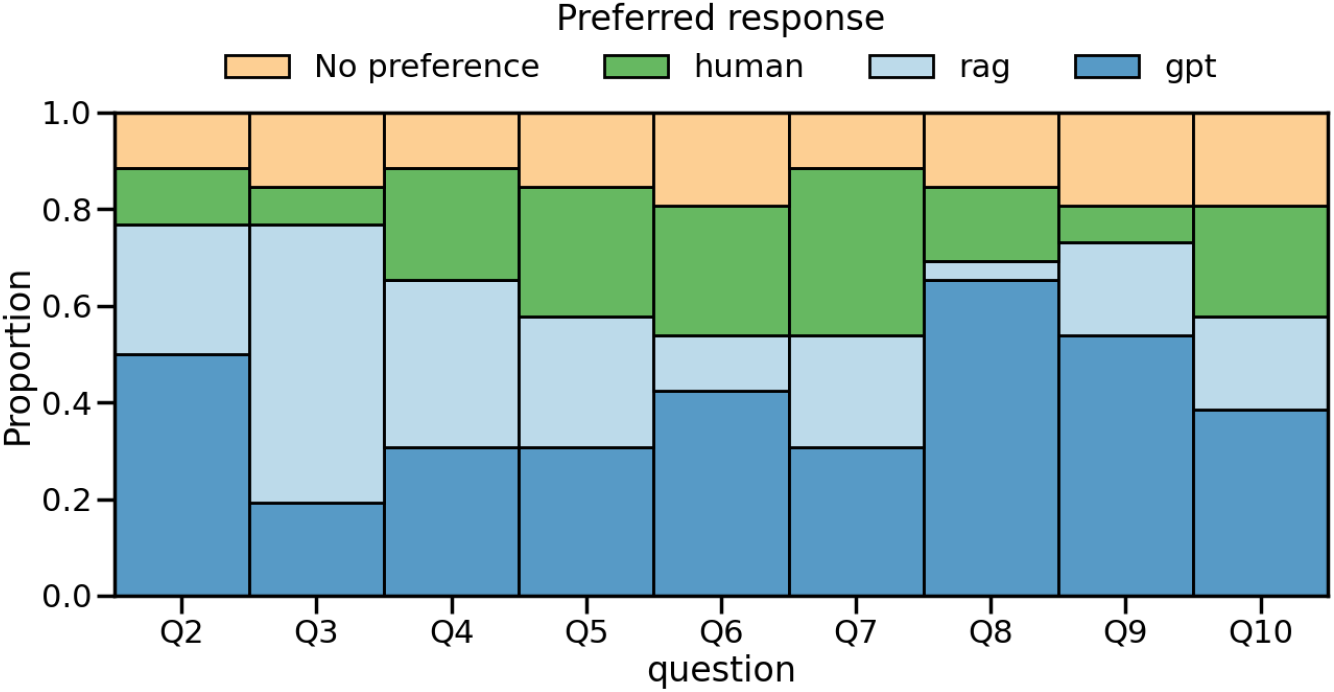
Proportion of votes for preferred response shown per question.

**Figure 3.**
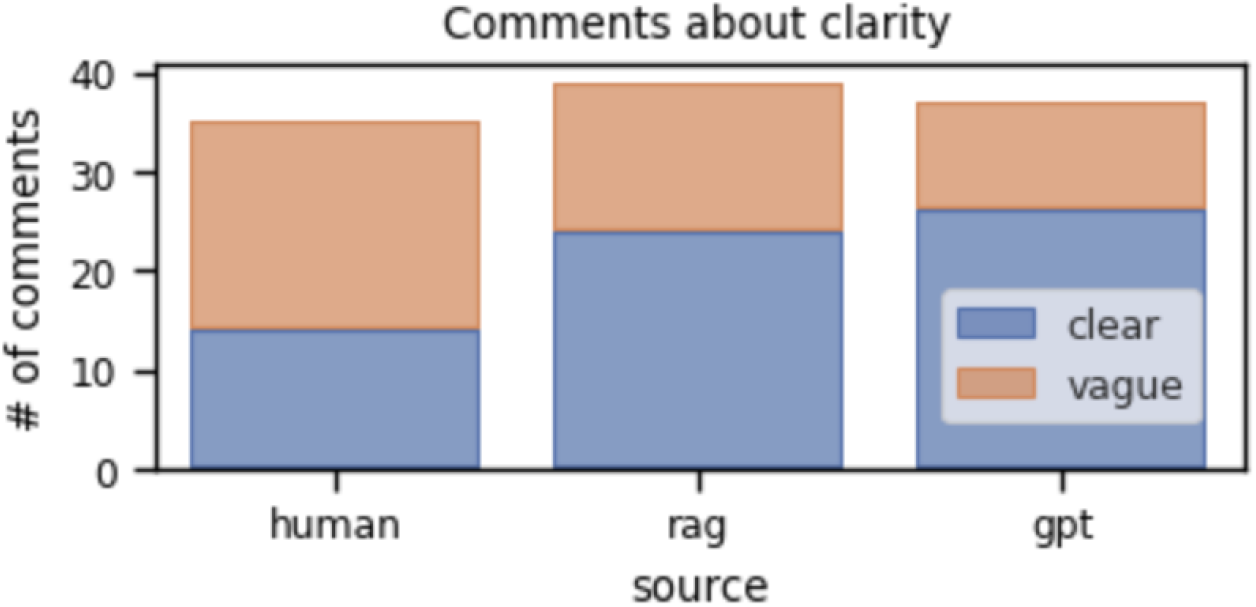
Count of comments annotated as mentioning either “clear” or “vague”.

Despite comments being indicated as optional, respondents provided 519 out of 702 possible comments (26 responses * 9 questions * 3 responses per question), with 17 respondents providing comments for at least 20 out of 27 total responses. As such, the free-text comments presented a rich resource for understanding patient preferences in detail. To this end, we conducted a thematic analysis aimed at understanding what attributes are most salient to patients, as well as their preferences along these attributes. Our initial code development phase identified five attributes that were mentioned across multiple responses: clarity, verbosity, certainty, personability, and numeracy. We separately annotated positive and negative mentions of each attribute, resulting in 10 unique labels annotated during the code application phase. We summarize the results of our thematic analysis in Table 2. Among these, clarity and verbosity constituted the most frequent annotations, respectively mentioned in 111 (21.3%) and 172 (33.1%) comments.

**Table 2.** Results of thematic analysis.

| Theme | Label | Description | Example | # tagged instances |
| --- | --- | --- | --- | --- |
| Clarity | **Clear** | Indicates response is clear or otherwise provides a good explanation | *Clear... easy to understand.* | 64 |
|  | **Vague** | Indicates response is vague, confusing, or poorly explained | *Answers the question but the explanation is not clear* | 47 |
| Verbosity | **Succinct** | Praise for succinct, direct answers OR criticism for overly verbose responses | *I like that this response is short and direct.* | 104 |
|  | **Verbose** | Praise for comprehensive, detailed responses OR criticism for responses that are too short or lacking in detail | *I liked this response because someone with two APOE4 genes will want to know in detail what they can do.* | 68 |
| Certainty | **Deterministic** | Preference for expressions of certainty | *I hate grey zones. I prefer black and white, true and not true. And empowering is not a term I would use because it's more like frustrating to me if someone says I'm in a grey area of life.* | 5 |
|  | **Probabilistic** | Preference for open-ended/probabilistic statements | *I like this and believe this is the MOST important message in this response -it's essential to remember that "risk" doesn't mean "certainty."* | 4 |
| Numeracy | **Quantitative** | Preference for data/numerical values | *This response is very specific in its percentages and breaks down the distribution of the combinations even more. I like that.* | 9 |
|  | **Verbal** | Preference for verbal descriptions | *Numbers and percentages sometimes are overwhelming and can be confusing whereas this answer is calming to me.* | 17 |
| Personability | **Personal** | Preference for responses that connect with reader personally | *I like it's compassionate perspective.* | 12 |
|  | **Impersonal** | Preference for objective responses that provide facts | *Don't like it talking to me personally* | 10 |

Qualitative analysis identified four themes: verbosity, certainty, empathy, and numeracy. Within each theme, we identify contrasting preferences. For example, within the theme of certainty, some comments express a preference for responses that convey high certainty, whereas other comments express a preference for more probabilistic statements. In general, we observed divergent preferences along these themes.

Patients tended to calibrate information presented in responses against their own prior knowledge about AD, and misalignment tended to erode trust. For example, when presented with the claim that “Some of our study participants with MCI might find that over time, their cognitive function […] improves back to what we’d expect with normal aging,” one patient commented, “*I never heard of anyone going back to normal*” and gave low numerical ratings to the response. Whereas statistical evidence can sometimes be helpful to support a claim, in some patients they can raise more questions than answers. One patient writes, “*I’m not sure I know enough to properly understand the results without knowing how this information was arrived at. Were these percentages gathered from people who had the disease and assumed that this applies to the whole world? I’m confused about how the results were calculated based on what was presented*.” Some patients also reported aversion toward overly technical descriptions, whereas other patients appreciated the level of detail. For example, regarding a description of AD pathology, one patient commented, “*too complicated for general populations*,” whereas the same described the same response as “*comprehensive and clear description*.”

## Discussion

Our proof-of-concept study provides insights into potential patient preferences regarding AI-generated responses for frequently asked questions related to AD and AD-related genetics. The results suggest that patients may rate AI-generated responses higher than human responses in terms of trustworthiness, relevance, and coherence. This suggests a potential patient preference for more comprehensive and consistent information delivery, often characterized by the verbose and detailed responses generated by LLMs^19^, although we also identified variability in patient preferences.

Free-text comments revealed verbosity and clarity to be salient attributes. Interestingly, while many patients expressed a preference for succinctness, as highlighted in Figure 3, more verbose responses typical of GPT were rated higher overall. This divergence underscores a trade-off between the need for detailed, comprehensive responses and the desire for concise communication. Prior studies have found similar trends, suggesting that detailed responses may evoke a greater sense of reliability and thoroughness, even if they are perceived as less efficient^11^.

Patients’ receptivity to AI-generated responses indicates a potential role for LLMs in supplementing or even enhancing traditional genetic counseling. However, our findings also highlight the heterogeneity in patient communication preferences. Some patients valued empathetic and personalized responses, while others preferred objective, fact-based explanations. Figure 2 shows the distribution of preferred responses across different questions, illustrating the context-dependent nature of these preferences. To meet these diverse needs, future AI systems must be flexible and capable of adapting their responses to suit individual patient preferences.

Our thematic analysis revealed additional dimensions influencing patient perceptions, including clarity, verbosity, certainty, empathy, and numeracy. Clarity, in particular, was a dominant factor, with AI responses often being described as either clear or vague, as illustrated in Figure 3. Patients tended to favor responses that provided actionable information and clear explanations, while responses perceived as overly technical or patronizing were less well-received. In particular, we believe it is important to calibrate materials to patients’ prior knowledge. While this study is an initial effort to explore patient preferences in answers to medical questions, future work is needed to confirm these findings in larger representative patient populations and should explore more broadly what constitutes a good answer to patients. Prior work, ours included, assumes that human-written answers are the “golden standard” for medical communication^20–22^. However, our results indicate that that might not necessarily be the case, as our respondents more often preferred machine-generated answers. Additionally, future work could explore how patients’ knowledge of a response being AI-generated affects their trust in a response.

Despite these promising findings, our study has several limitations. First, this is a small human evaluation and larger studies in representative patient populations are needed to confirm these findings. Demographics of the participants were not collected and data was not able to be analyzed to identify correlation between participant preferences and demographics such as education level, socioeconomic status, gender, age, etc. Additionally, respondents were already familiar with genetic counseling, which may have influenced their evaluations and critiques based on preexisting knowledge. This familiarity could have introduced a bias that favors or discredits certain response styles. Additionally, we did not collect expert annotations to compare with patient feedback, which limits our ability to assess clinical accuracy and appropriateness.

Another limitation includes differences in the way that the genetic counselors and GPT were prompted, which may contribute to differences in responses generated. Human counselors were asked to craft responses to FAQ-style questions, resulting in more direct and succinct responses, while AI systems were prompted to “respond like a genetic counselor,” leading to a more verbose explanatory style. Genetic counselors typically use empathy and personalize counseling discussions when interacting directly with patients, but preferred direct and succinct responses in the FAQ setting. This distinction also highlights a shift in the native communication domains of clinicians and AI – whereas patient-provider interactions occur primarily by speech, with writing used mostly for documentation, LLMs are optimized for text-only communication. The text-based evaluation setting may constrain the expressiveness of human clinicians. Overall, these gaps may limit the generalizability of our results to the patient-provider interaction setting, and future work should address AI systems deployed in more realistic clinical scenarios.

## Conclusion

In this pilot study, we found that AI-generated responses were well received by patient and could show promise in enhancing patient-centered communication. Ongoing efforts may benefit from tailoring responses to meet individual preferences, preventing hallucinations, and fostering open-ended patient evaluations. This approach will ensure that AI systems can be seamlessly integrated into genetic counseling and other healthcare domains to better provide equitable, effective, and personalized care.

## Data Availability

All data produced in the present study are available upon reasonable request to the authors

