## Supplement S1 for "Patient-centered Evaluation of AI Answers to Genetic Counseling Questions"

Q1: Do we know of other genes that pose a risk to dementia?

RAG

While APOE is the most well-known and best-studied genetic risk factor for late-onset Alzheimer's disease, there are other genes that have been identified to be associated with Alzheimer's risk. However, the impact of these genes on overall risk is generally much smaller compared to APOE.

When we discuss the scope of risk, it's important to understand that most other genes contribute a relatively minor increase or decrease in Alzheimer's risk. The combined effects of these other genes may influence one's overall risk, but they don't confer the same level of risk as the APOE E4 variant.

Considering the broader effects of risk, it's essential to remember that genetics is just one piece of the puzzle. An individual's overall risk is shaped by a combination of genetic factors, environmental influences, lifestyle choices, and health conditions. Even with the presence of multiple risk genes, other factors like maintaining good cardiovascular health and managing conditions like diabetes can play a pivotal role in modifying the risk.

Human

Yes, other genes besides APOE are associated with increased risk to develop dementia. However, these genes are relatively rare and typically found in families with a strong history of dementia and/or other related diseases. If you have questions or concerns about other genes increasing risk for dementia in your family, a neurology genetic counselor may assess your personal and family history and offer testing for additional genes, if indicated.

GPT-4

There is a growing understanding in the field of genetics regarding the complexity of dementia and its hereditary components. While the APOE ε4 allele is the most well-known genetic risk factor for late-onset Alzheimer's disease, which is a form of dementia, there are indeed other genes that have been associated with an increased risk for various types of dementia.

Some genes are directly linked to rare, early-onset forms of Alzheimer's disease, including PSEN1, PSEN2, and APP. Mutations in these genes can lead to disease onset in an individual's 30s, 40s, or 50s.

Furthermore, research has identified numerous other genes that can modulate the risk for developing Alzheimer's disease and other dementias to a lesser extent than APOE. These genes may influence processes like inflammation, lipid metabolism, and tau protein buildup.

It's also important to note that for many people, the risk of developing dementia is a combination of genetic, environmental, and lifestyle factors. Genetics can provide some insight into risk, but they don't paint the entire picture. Regular medical check-ups, maintaining a healthy lifestyle, and staying mentally active are crucial for overall brain health.

If you or a loved one are concerned about your genetic risk for dementia, it might be beneficial to seek out genetic testing and counseling to gain a more comprehensive understanding of your personal risk and potential preventive strategies.

Q2: What's the difference between dementia, mild cognitive impairment, and Alzheimer's disease?

RAG

You know how as we age, we might notice changes in our abilities to remember names or misplace our keys? That's a part of the normal aging process. But, imagine a friend from our study group who starts to have slightly more pronounced memory challenges, maybe forgetting the topic of a conversation they just had. That's what we call mild cognitive impairment, or MCI. It’s like being in between normal aging and something more severe. Some of our study participants with MCI might find that over time, their cognitive function either stays the same, improves back to what we'd expect with normal aging, or it could progress further.

Now, imagine another friend from our group who not only forgets conversations but also struggles with daily activities, like getting dressed or preparing a meal. That's what we refer to as dementia. Dementia isn’t a specific disease itself, but a broad term describing a set of symptoms that significantly impact daily living.

Among the various causes of dementia, Alzheimer's disease is the most common. So, if we were to picture dementia as an umbrella, Alzheimer's disease would be one of the main spokes holding it up.

To sum it up: MCI is like a gray zone, a bit beyond normal aging but not as severe as dementia. Dementia, including Alzheimer’s, affects daily life much more profoundly. Since you've come to learn about the APOE E4 genes, it shows you're already informed about the risk factors. This knowledge can be empowering and help you better understand where you might fit in the larger picture of our study and the community of participants.

Human

Mild cognitive impairment, also called MCI, refers to a small but noticeable and measurable decline in cognition. Individuals with MCI may remain stable, return to aging normally, or progress to dementia. Dementia is a general term for a decline in mental ability severe enough to interfere with daily life. There are many different causes of dementia, but Alzheimer’s disease is the most common. Alzheimer’s disease is a progressive disease that causes decline in memory, thinking, and behavior over time.

GPT-4

Dementia is a general term that describes a group of symptoms related to memory loss, thinking, and social abilities severe enough to interfere with daily life. It is not a specific disease in itself but rather a clinical syndrome, meaning a group of symptoms that occur together. There are various causes of dementia, with Alzheimer's disease being the most common cause.

Mild cognitive impairment (MCI) can be seen as an intermediate stage between the expected cognitive decline of normal aging and the more pronounced decline of dementia. People with MCI have noticeable memory problems or other cognitive issues that are more significant than typical age-related changes, but these issues are not severe enough to interfere significantly with daily life. Not everyone with MCI will progress to dementia, but they are at an increased risk.

Alzheimer's disease, on the other hand, is a specific type of dementia. It's a progressive neurological disorder that leads to memory loss and cognitive decline. It is characterized by the abnormal buildup of proteins in the brain, leading to the death of brain cells. While Alzheimer's disease is the most common cause of dementia, it's essential to remember that not all dementia is due to Alzheimer's. There are other causes, such as vascular dementia, Lewy body dementia, and frontotemporal dementia, among others.

Q3: Percentage-wise, how many people have each or fallen to each of these categories of E2, E3 and E4?

GPT-4

The APOE gene plays a role in fat metabolism and has been associated with the risk of Alzheimer's disease, among other conditions.

E3 is the most common allele globally, with the majority of people inheriting two copies of E3 (one from each parent). In many populations, over 50% of individuals have this genotype (E3/E3).

E4 is associated with an increased risk of developing Alzheimer's disease. If you inherit one copy of the E4 allele, your risk is increased, and if you inherit two copies, your risk is even higher. Around 20-25% of the general population carries at least one E4 allele.

E2 is the least common of the three alleles and is associated with a decreased risk of developing Alzheimer's disease. Only a small fraction of the population carries the E2 allele, with estimates suggesting less than 10% have at least one copy.

Human

Each person inherits an APOE allele from each biological parent, meaning people can have one of six possible combinations: e2/e2, e2/e3, e2/e4, e3/e3, e3/e4, and e4/e4. Among these APOE alleles, e3 is the most common. About 60% of the population has the e3/e3 result. About 20-25% have at least one copy of e4 and 2-5% have two copies of e4. Only about 7% of individuals have the e2 allele.

RAG

When we look at the ApoE alleles, E3 is the most common, with a vast majority of people carrying it. E2 is on the rarer side, which means fewer people have it. E4 is somewhere in the middle. To give you a clearer picture, about 20% of the general population has one E4 allele. Now, when we talk about having two E4 alleles, like you do, only about 2-4% of the population falls into that category.

It's important to note that these percentages just give an overview of the genetic landscape; they don't predict outcomes for individuals. Think of it like this: just because you're part of that 2-4% doesn't mean you're on a set path towards a particular condition. Genes, including E4, are merely one of many factors influencing our health. The presence of E4 does indicate a higher risk for certain conditions, specifically mild cognitive impairment or Alzheimer's-related dementia. But it's essential to remember that "risk" doesn't mean "certainty." It's similar to how some people might have a family history of a condition but never develop it themselves.

I often remind individuals that genetic testing, like this, gives us a risk assessment. It's like looking at the weather forecast; a higher chance of rain doesn’t always mean it will rain. Similarly, carrying the E4 allele increases the likelihood but doesn't determine if one will develop symptoms. The human body is complex, and many factors, both genetic and environmental, play a role in our overall health.

Q4: How common is the E4 type of APOE?

GPT-4

The APOE gene, which is associated with cholesterol metabolism and the risk of developing Alzheimer's disease, has several variants. Of these, the ε4 variant (often simply referred to as E4) is of particular interest because it's linked to an increased risk for late-onset Alzheimer's disease.

The prevalence of the ε4 allele varies widely among different populations. In many populations, around 10-15% of individuals carry at least one ε4 allele. However, it's essential to highlight that having the ε4 allele doesn't guarantee someone will develop Alzheimer's disease; it merely increases the risk. Conversely, not having the ε4 allele doesn't mean someone won't develop the disease.

The frequency can be higher or lower depending on specific populations and ethnic backgrounds. For instance, certain populations might have a much lower prevalence of the ε4 allele, while others could have a higher prevalence.

If you or a loved one are considering genetic testing related to Alzheimer's risk or any other health concerns, it's crucial to understand the implications and limitations of such results. It's always recommended to work with a genetic counselor or other medical professional who can provide guidance tailored to individual circumstances.

Human

About 25% of the general population carries at least one copy of the e4 type of APOE.

RAG

In terms of the broader population, E4 is a fairly common version of the APOE gene. E3 is the most prevalent type, while E2 is the least common. To put it in perspective for you, about 20% of individuals in the general population have one copy of the E4 allele, much like many others participating in the study. A smaller group, around 2-4%, has two copies of the E4 allele, just like you. It's essential to remember that while the E4 allele does increase the risk for mild cognitive impairment or Alzheimer's-related dementia, it's not a definitive marker. Many people with the E4 allele never develop any symptoms. There's still much we're learning about the complexities of how genes and environmental factors interact. You're not alone in this, and many participants in the study are on this journey of understanding alongside you.

Q5: What age is considered early for early onset Alzheimer's?

RAG

Early-onset Alzheimer's typically refers to individuals who develop symptoms before the age of 65. Now, considering the data we discussed, many people, even with two copies of the E4 variant, like you, don't show symptoms until between ages 65-85. That's a range in which the risk is highest. However, it's essential to understand that these numbers are based on large studies and there's a level of uncertainty. Just as 30-55 out of 100 people with your APOE result might develop symptoms, 45-70 might not. It's also worth noting that those lifestyle changes we talked about have been shown to potentially reduce the risk, highlighting how other factors come into play. We're all different, and individual experiences may vary.

GPT-4

Most individuals with early-onset Alzheimer's begin experiencing symptoms in their 50s, but it can also manifest in one's 40s or even as early as the 30s in some rare cases. This form of the disease is less common than late-onset Alzheimer's, which typically appears after the age of 65. If there are concerns about memory or cognitive changes at a younger age, it's essential to consult with a medical professional. Genetic factors can play a significant role in early-onset Alzheimer's, so understanding family history and potentially considering genetic testing can provide valuable information about risks and possible interventions.

Human

Alzheimer’s disease is considered “early-onset” when the disease develops before age 65.

Q6: What is the risk for Alzheimer's with each type of APOE result?

RAG

With the specific E4 E4 makeup that you have, the estimated risk for developing Alzheimer's disease by age 85 falls between 30-55%. This means there's a significant chance, between 45-70%, that you may not develop Alzheimer's by that age. For those with the E3 E4 combination, the risk is approximately 20-25%. Other APOE combinations generally have a risk of 10-15% or even less.

However, it's essential to keep in mind that these risk estimations are not absolutes. They don't account for lifestyle, environmental factors, or other potential genetic influences that may either increase or decrease the risk. It's a complex interplay of multiple factors. Knowing your genetic information can certainly guide prevention and wellness efforts, but it's just one part of the broader picture of your health.

RAG

Everyone has some risk of developing Alzheimer’s disease.  The most common APOE result is e3/e3, which is associated with a 10-15% risk of developing Alzheimer’s disease by age 85. The e3 type of APOE is neutral, it does not increase or decrease the risk of Alzheimer’s disease.  When someone has one or two copies of the e4 type, the risk can increase.  The e3/e4 result has a 20-25% risk and the e4/e4 result has a 30-55% risk by age 85. The e2 type of APOE is more rare, which means we have less data about how it impacts the lifetime risk of Alzheimer’s disease.  The data we do have suggests that the e2 type of APOE reduces the risk for AD.  The risks for the e2/e2 and e2/e3 results may be slightly lower than the e3/e3 result, while the e2/e4 result might be slightly higher or slightly lower than the e3/e3 result.

GPT-4

The APOE gene plays a significant role in the risk of developing late-onset Alzheimer's disease, and it has three common alleles: ε2, ε3, and ε4.

APOE ε3 is considered the "neutral" allele, meaning that individuals with two ε3 alleles (ε3/ε3) have an average risk of developing Alzheimer's disease relative to the general population.

APOE ε4 is associated with an increased risk. Having one ε4 allele (ε3/ε4) increases your risk compared to those with ε3/ε3, and having two ε4 alleles (ε4/ε4) further increases this risk. It's worth noting that not everyone with one or even two ε4 alleles will develop Alzheimer's disease; it merely increases the likelihood. Many other factors, including other genetic, environmental, and lifestyle factors, come into play.

Conversely, APOE ε2 is considered protective. Individuals with at least one ε2 allele, such as ε2/ε3 or ε2/ε2, tend to have a lower risk of developing Alzheimer's disease compared to those with ε3/ε3. However, the presence of an ε2 allele doesn't guarantee protection; it only reduces the risk.

Remember that while APOE is a significant genetic risk factor for Alzheimer's, it is not deterministic. Many individuals with ε4 alleles never develop the disease, and some without ε4 alleles do. It's a complex interplay of genetics, environment, and lifestyle. If you or a loved one have concerns about your APOE status and risk for Alzheimer's, consulting with a medical professional can provide a clearer understanding and guidance on potential preventive measures.

Q7: Will my children have the same APOE result as me?

GPT-4

Each person has two copies of the APOE gene, one inherited from each parent. The APOE gene has three main variants (or alleles): ε2, ε3, and ε4.

If you've had genetic testing that provided your APOE status, you'll have two of these alleles. For example, you might have an ε3/ε4 combination or an ε2/ε3 combination.

Your children will inherit one APOE allele from you and one from their other parent. So, while they will inherit one of your two alleles, they may not necessarily have the same combination as you, depending on what alleles their other parent contributes.

For instance, if you have an ε3/ε4 combination and your partner has an ε3/ε3 combination, your child could inherit the ε3 allele from both of you (ending up with an ε3/ε3 combination) or could inherit the ε4 allele from you and the ε3 allele from your partner (resulting in an ε3/ε4 combination).

In summary, your children will inherit one of your APOE alleles, but their exact APOE result will also depend on the genetic contribution from their other parent. If you have concerns or questions about the potential genetic risks for your children, a detailed discussion with a genetic counselor can provide clarity and guidance tailored to your family's specific situation.

Human

Your children may or may not have the same APOE result as you.  This is because your children inherit one copy of APOE from you, and their second copy of APOE comes from their other parent.  For example, if you have two copies of the e3 type of APOE, you will always pass on one copy of e3 to your children.  But their final result will be determined by what type of APOE they inherit from their other parent.

RAG

Based on your genetic profile, your children will definitely inherit one copy of the E4 variant from you. However, without knowing the genetic status of their other parent, we cannot determine what the second copy of the ApoE gene will be for your children.

It's important to understand that while inheriting an E4 variant does increase their risk for mild cognitive impairment or Alzheimer's disease, it doesn't guarantee it. If we look at the bigger picture, individuals with two copies of the E4 variant have a 30-55% risk of developing symptoms by age 85. This means there's a 45-70% chance they may not develop Alzheimer's. Various factors influence the onset and progression of the disease.

Discussing your results with your children when they're ready can be crucial, as it might have broader implications for their health choices and future planning. While the information brings a certain level of uncertainty, understanding the scope of the risk can help make informed decisions.

Q8: Are there ways to lower my risk of developing Alzheimer's disease?

RAG

It's essential to understand that the 30-55% risk estimate for developing mild cognitive impairment or Alzheimer's disease by age 85 is based mainly on a specific large study, among other data. This estimate is just that—an estimate. It's not a guarantee that someone will develop Alzheimer's, and there are various factors, both genetic and environmental, that can influence one's risk.

With two APOE E4 genes, while your genetic predisposition might be higher, there are actionable steps you can take to possibly reduce your risk. Engaging in regular exercise, maintaining a heart-healthy diet, managing stress, and if you smoke, quitting, are all strategies grounded in research aimed at promoting healthy brain aging. Additionally, addressing conditions like cardiovascular disease, diabetes, and high blood pressure can be beneficial, as these can push one closer to the higher end of the risk spectrum. Being proactive in these areas can serve as a defense against the potential onset of cognitive impairment or Alzheimer's disease.

Human

Lifestyle changes that are healthy for the heart are generally considered healthy for brain aging. These lifestyle changes may include: exercise, eating a heart-healthy diet, reducing stress, and quitting smoking. It is also recommended to engage in social and mentally stimulating activities. Although none of these activities has been proven to prevent mild cognitive impairment or dementia due to Alzheimer’s disease, they may have other health benefits (such as reducing risk for cancer and heart disease).

GPT-4

While genetic factors play a role in the risk of developing Alzheimer's disease, especially in its early-onset form, there are various modifiable factors and lifestyle choices that can influence your risk. By addressing these, it's possible to reduce the likelihood of developing Alzheimer's or delay its onset.

Physical activity is one of the most beneficial strategies. Regular exercise helps maintain good blood flow to the brain and encourages the growth of new brain cells. This doesn't necessarily mean vigorous workouts; even walking daily can make a difference.

Diet also plays a significant role in brain health. Diets rich in fruits, vegetables, whole grains, fish, and lean proteins, such as the Mediterranean or DASH (Dietary Approaches to Stop Hypertension) diets, have been associated with a reduced risk of cognitive decline.

Maintaining good heart health is crucial too. Conditions like hypertension, diabetes, and high cholesterol can increase the risk of Alzheimer's disease. So managing these conditions through medication, diet, and lifestyle can have a protective effect on the brain.

Mental stimulation is another essential aspect. Engaging in activities that challenge the brain, like reading, solving puzzles, or learning a new skill, can strengthen brain cells and the connections between them. Social engagement, such as participating in group activities or maintaining close personal relationships, has also been linked to a lower risk of cognitive decline.

Finally, it's vital to monitor and manage your mental health. Conditions like depression, stress, and sleep disturbances can impact cognitive health. Seeking treatment when needed and adopting stress-reducing practices can be beneficial.

While these strategies can help reduce the risk or delay the onset of Alzheimer's disease, they don't guarantee prevention. However, adopting a combination of these healthy behaviors can promote overall well-being and cognitive health as you age. If you have specific concerns or a family history of Alzheimer's, it might be helpful to discuss personalized strategies with a medical professional.

Q9: How much memory loss is equivalent to just aging normally?

Human

This is a difficult question to answer. While some occasional forgetfulness can be a normal part of aging, it would be important to consult with a doctor if you are noticing these difficulties occurring more often, or if they are interfering with normal daily life.

GPT-4

It's entirely normal for some memory changes to occur as part of the natural aging process.

As we age, you might notice that it takes longer to learn new things, you don't retain information as easily as you once did, or you might misplace objects like your keys or glasses more often. It's also not uncommon to occasionally forget a person's name or an appointment but remember it later.

However, the kind of memory loss associated with normal aging does not severely disrupt daily life. It's often transient and not progressive. For example, forgetting the name of an acquaintance is typical, but forgetting the name of a close family member or the function of everyday items, like what a key is used for, is not.

It's crucial to differentiate between these typical age-related changes and more severe memory problems that could be symptoms of a more serious condition, like mild cognitive impairment, Alzheimer's disease, or another form of dementia.

If memory problems become concerning or start to interfere with everyday activities, it's essential to seek a medical evaluation. Other factors, such as medication side effects, thyroid issues, or vitamin deficiencies, can also impact memory, and addressing these underlying causes can improve cognitive function.

Always remember that while some memory changes are a normal part of aging, staying mentally active, engaging in regular physical activity, maintaining social connections, eating a balanced diet, and managing chronic conditions can all help support brain health as one ages.

RAG

When we talk about the memory loss associated with normal aging, we're generally referring to occasional forgetfulness, like misplacing keys or forgetting an acquaintance's name. This kind of memory change is common and expected as people age. It's different from the more significant cognitive decline seen in conditions like mild cognitive impairment or Alzheimer's disease.

Q10: What does Alzheimer's look like inside the brain?

RAG

Alzheimer's disease is characterized by the accumulation of abnormal protein deposits, mainly amyloid plaques and tau tangles, which interfere with neuronal function. Over time, these deposits cause neurons to lose their connections and eventually die, leading to memory loss and cognitive decline. Picture it like a garden where some plants start to wither and the pathways between them become obstructed. But remember, not every garden with these signs will end up in disarray. Many gardens, or in this case brains, remain resilient.

Human

When a brain is affected by Alzheimer’s, certain areas of the brain will be shrink (become smaller).  Doctors often call this shrinking “atrophy” and sometimes it can be seen using a brain scan (for example, a MRI).  If you were able to look inside the affected brain cells, you would see build-up of two specific proteins.  One of these proteins is called beta-amyloid and the other is called tau.  Some memory clinics are now able to detect these proteins using a special kind of brain scan, which helps doctors more accurately diagnosis Alzheimer’s disease.

GPT-4

Alzheimer's disease results in distinct changes within the brain at both the microscopic and macroscopic levels, which are characteristic of the disease.

At a microscopic level, Alzheimer's is marked by the accumulation of two types of abnormal protein structures: amyloid plaques and tau tangles. Amyloid plaques are sticky clumps of protein fragments that accumulate outside of nerve cells, or neurons. These plaques may disrupt cell function and trigger inflammation, leading to neuron damage. Tau tangles, on the other hand, are twisted fibers that form inside neurons. They result from the abnormal buildup and twisting of a protein called tau. These tangles can block the neuron's transport system, impeding nutrient and other essential molecule movement, leading to cell death.

On a more macroscopic level, as Alzheimer's disease progresses, significant brain atrophy or shrinkage occurs. This is especially noticeable in the brain's hippocampus, an area crucial for memory formation. As more neurons die due to the disease, the affected brain regions start to shrink, leading to a decline in cognitive functions. Over time, this shrinkage becomes more widespread, which is often evident when comparing brain scans of healthy individuals to those with advanced Alzheimer's.

Fluid-filled spaces in the brain called ventricles also enlarge as the brain tissue decreases. Moreover, the disease may disrupt the usual activity and connectivity between different brain regions, impairing communication between neurons.

Understanding the changes that Alzheimer's disease causes in the brain is crucial for its diagnosis and ongoing research into potential treatments. If you or a loved one have concerns about Alzheimer's or are considering pursuing more detailed diagnostic procedures, it's essential to consult with medical professionals familiar with the disease.
